# Mast cells differentiated in synovial fluid and resident in osteophytes exalt the inflammatory pathology of osteoarthritis

**DOI:** 10.1101/2021.11.14.21266312

**Authors:** Priya Kulkarni, Abhay Harsulkar, Anne-Grete Märtson, Siim Suutre, Aare Märtson, Sulev Koks

**Affiliations:** Department of Pathophysiology, Institute of Biomedicine and Translational medicine, University of Tartu, Ravila 19, Tartu- 50411, Estonia; Department of Pharmaceutical Biotechnology, Poona College of Pharmacy, Bharati Vidyapeeth University, Erandwane, Pune – 411038, India; Department of Pharmacology and Therapeutics, University of Liverpool, UK; Department of Anatomy, Institute of Biomedicine and Translational medicine, University of Tartu, Ravila 19, Tartu- 50411, Estonia; Department of Traumatology and Orthopaedics, Institute of Clinical Medicine, University of Tartu, L Puusepa 8, Tartu- 51014, Estonia; Clinic of Traumatology and Orthopaedics, Tartu University Hospital, L Puusepa 8, Tartu- 51014, Estonia; Perron Institute for Neurological and Translational Science, Nedlands, 6009, Western Australia; Centre for Molecular Medicine and Innovative Therapeutics, Murdoch University, Murdoch, 6150,Western Australia

**Author notes:** **Corresponding Author –** Sulev Koks –. **Email –** Priya Kulkarni –; Abhay Harsulkar –; Anne-Grete Märtson –,; Siim Suutre –; Aare Märtson –. (**Note**: The last two authors contributed equally for the present manuscript).

## Abstract

Osteophytes are a prominent feature of osteoarthritis (OA) pathology. RNA-seq of osteophytes revealed patterns corresponding to active ECM re-modulation and participation of mast cells. The cells recruitment and their activity status were confirmed by anti-TPSAB1 and anti-FC epsilon RI antibodies in immunohistochemistry. Besides subchondral bone, which is a logical yet unproven route for the cells deployment into osteophytes, the authors propose that OA synovial fluid (SF) is necessary and sufficient for maturation of mast cell precursors (MCPs) in this channel. The authors present evidences to support their claim in the form of IHC, proteomics analysis of SF samples and in vitro cell differentiation assay, wherein human monocytes (ThP1) and hematopoietic stem cells (HSCs) showed differentiation in HLA-DR+/CD206+ and FCERI+ phenotype respectively after 9 days of SF treatment. These observations expound osteophytes and resident mast cells as yet unexplained functional epicentre in OA pathology.

## Introduction

Osteophytes, commonly known as ‘bone spurs’, are frequently seen in synovial joints like spine, hand and knee. These are marginal ectopic formation of osteo-cartilaginous metaplastic tissue mostly at the junction of periosteum and synovium that appear to merge with or overgrown with the original articular cartilage. Although, osteophytes do not necessarily warrant any clinical intervention, depending on the position they can cause nerve compression in spine and more friction in the knee joints that lead to crepitus, discomfort and pain and may require surgical resection [**1**]. Clinically, osteophytes define structural progression of osteoarthritis (OA) along with the other clinical signs including joint space narrowing, subchondral sclerosis and the cartilage defects.

A process of osteophytes formation is known as ‘osteophytosis’. During osteophytosis, mesenchymal stem cells at the joint differentiate into chondrocytes showcasing different signatures of chondrogenesis. Further, centrally placed well differentiated hypertrophic chondrocytes subsequently undergo osteoblastogenesis and facilitate the growth of osteophytes. This entire process is similar to endo-chondral ossification that takes place in the foetal growth plate formation [**1**]. A fully grown and mature osteophyte is a bony outgrowth with a cartilage cap and minuscule of bone marrow [1]. In the recent study, Roelof et al., 2020 reported PDGFRα-expressing GDF5-decendant cells, including SOX9 progenitor cells and PRG4-expressing cells from synovial lining contribute to osteophytes formation. In the osteophytes, these cells are the source of hybrid skeletal cells and chondrocytes, respectively [**2**].

Osteophytosis is primarily attributed to synovial inflammation and particularly to macrophages, which are harbingers of synovitis. Macrophages play an essential role in a release of transforming growth factor-β (TGF-β), bone morphogenic protein-2 (BMP-2) and BMP-4, the key players involved in the osteophyte’s formation process and experimental depletion of these cells cause a significant reduction in the osteophytes, despite of a high dose intra-articulate injections of TGF-β, as shown in the murine model [**3**]. *In vivo* studies showed that inhibition of TGF-β or over-expressing TGF-β antagonist prevents osteophyte formation [**4, 5**].

Osteophytes have been investigated with high interest for their chondrogenic potential to decipher *in vivo* mechanism of cartilage regeneration. However, analysis of a structural composition of newly formed matrix revealed differences from a normal make-up of high-quality hyaline articular cartilage [**6**]. In another view, osteophytosis is proposed as repair mechanism in degenerating joints since the osteophytes at right place may help to stabilize the joint. This is because animal studies showed that surgical removal of the osteophytes can destabilize the joints [**7**]. However, random appearance of the osteophytes contradicts this purview. Further their association with symptoms like restricted joint movement, discomfort and pain put them in a pathological perspective and questions their biomechanical benefits. Osteophytes therefore are a pathological feature in OA which are worthy of investigation in molecular and cellular context.

Transcriptome analysis of a fully grown osteophyte is never attempted to gain deeper insight of pathophysiology associated with them. The authors performed a genome wide transcriptome analysis of six paired samples, which included osteophytes and non-osteophytic bony tissue (as a control) obtained from medial plateau of tibia in OA patients. This investigation revealed patterns corresponding to extra-cellular matrix (ECM) re-modulation, tissue damage and recruitment and activation of immune cells like, mast cells in osteophyte samples. Involvement of mast cells in OA pathology has been indicated by several studies [**8, 9, 10**]. Presence of mast cells and their degranulation products were reported in OA synovium and SFs [**8, 10**]. Based on the transcriptomics results, the authors further explored an association between osteophytes and mast cells, which is yet an untouched topic. They provide evidences for mast cell presence in osteophytes of OA joints and demonstrate their maturation mediated by synovial fluid (SF) in temporal fashion with progression of the disease.

## Results

### Differential transcripts between osteophytes and control specimens

After statistical analysis, 595 genes were differentially expressed between osteophytes and non-osteophytic controls (logFC ≥ 2 and logFC ≤ -2). Out of 595 genes, 322 genes were found up-regulated (LogFC≥ 2), while 273 were down-regulated (LogFC≤ -2) (**Figure 1 a** and **b, Figure 2 c**). Considering the scope of the hypothesis only up-regulated genes are described and discussed. In order to find out highly significant up-regulated genes, K-means cluster analysis was performed, wherein optimum number of clusters was determined by the elbow method. For up-regulated genes, optimal K was 7. Here, cluster numbers 1, 5 and 7 showed high significance (*P*< 0.001) and were considered for further analysis (**Figure 2 a** and **b**). Among 322 up-regulated genes, 87 genes were found with a marked increase in their expression levels (*P*< 0.001). All 87 up-regulated genes and 22 down-regulated genes are listed with their *P*-value and fold change in Supplementary Table 1 (**ST 1**).

**Figure 1.**
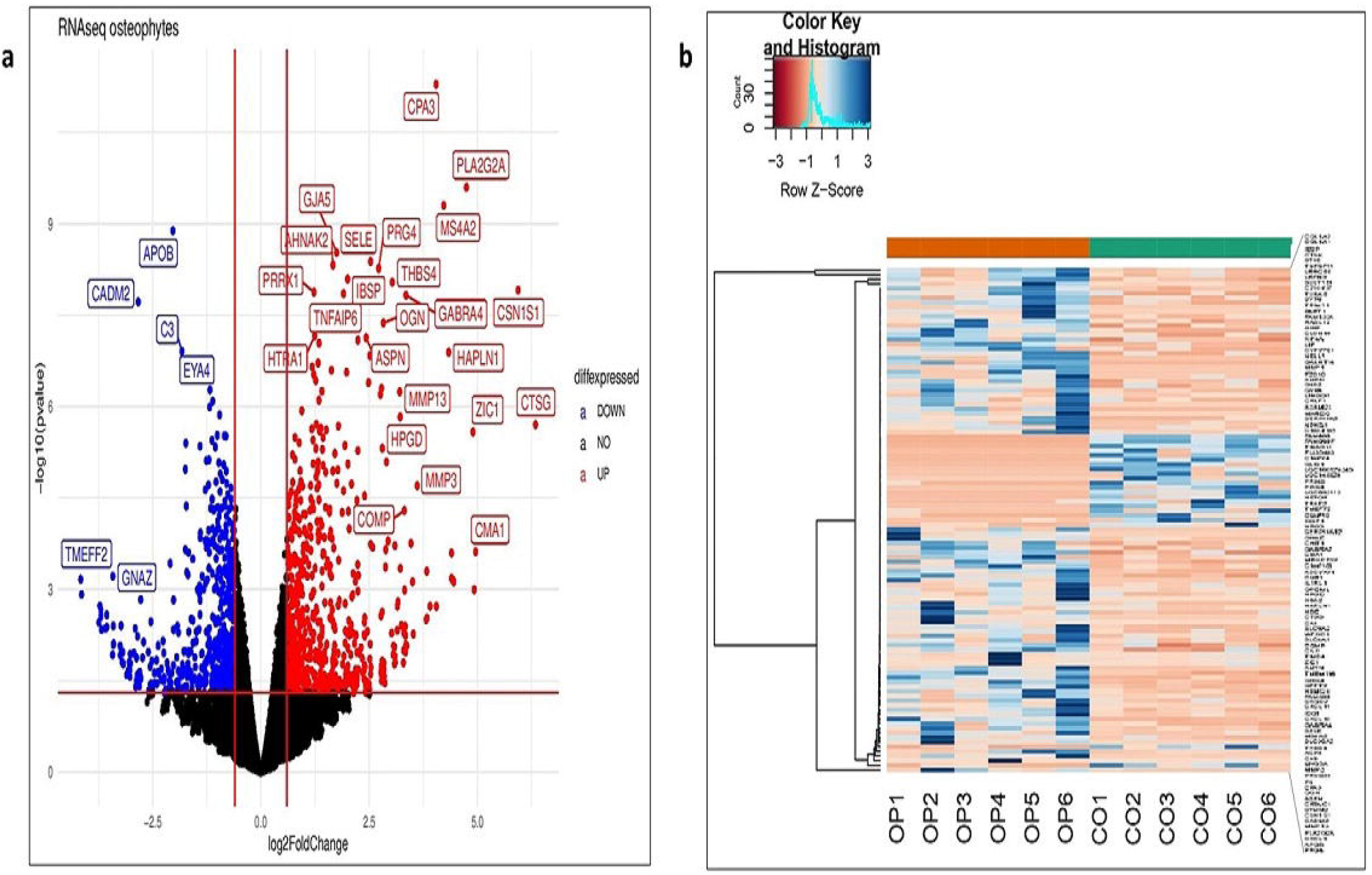
demonstrate an overview of differentially expressed genes in transcriptome analysis performed using osteophyte samples and non-osteophytic control tissue obtained from six knee OA patients (n=595) **a.** Volcano plot of the differentially expressed genes. Here, black dots represent insignificant genes, while blue and red dots represent down-regulated and up-regulated genes, respectively. The plot is generated using LogFC values of the expressed genes. CPA3, SELE, MS4A2/FCERI, CMA1, IL1RL1, COL1A1, COL1A2, MMP-1, MMP-3 and MMP-13 are among the significantly upregulated genes **b.** Histogram of the highly significant genes, which were obtained after K-means cluster analysis

**Figure 2.**
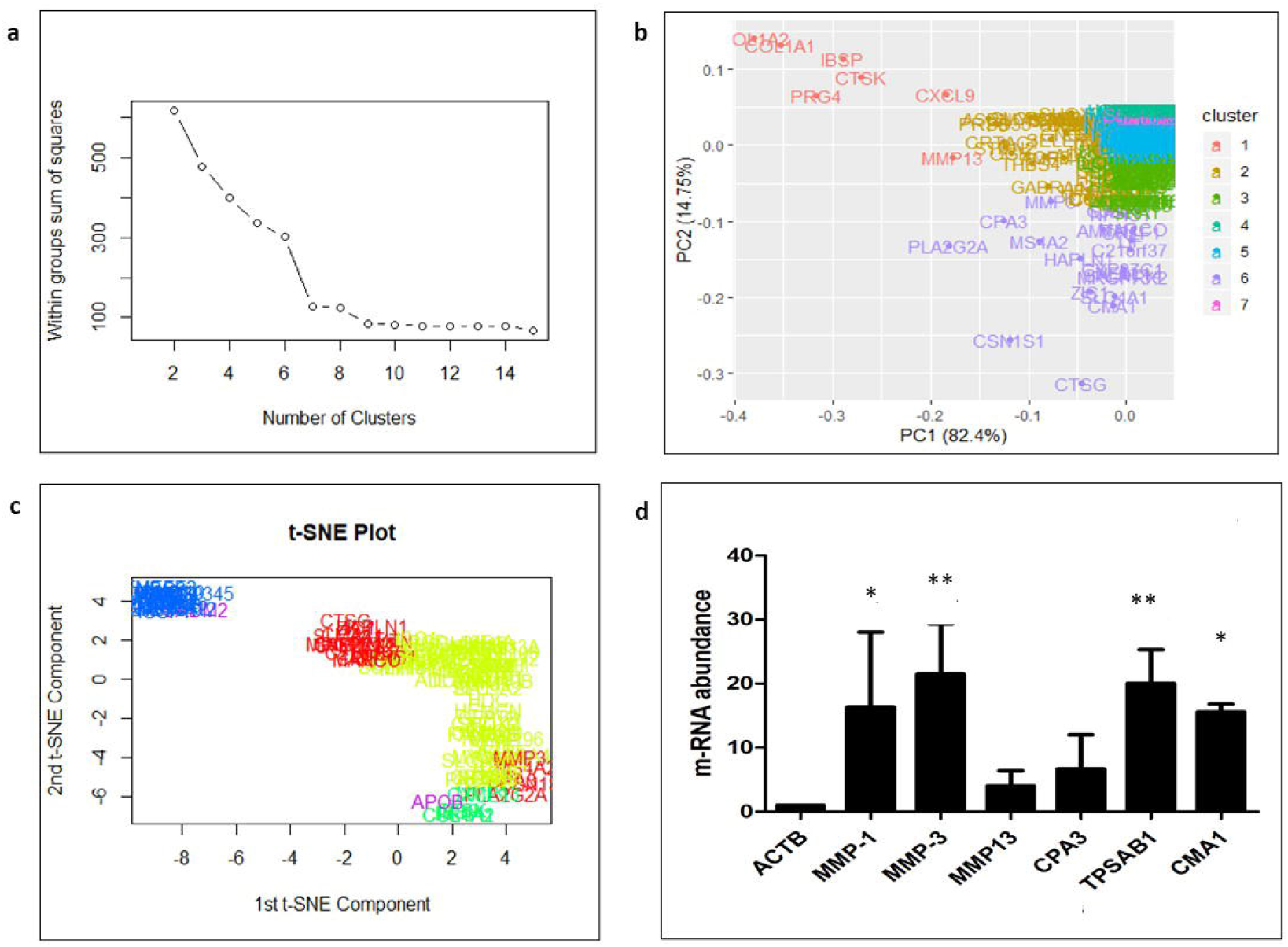
Cluster analysis of differential gene expression between osteophyte and control samples **a.** Denotes elbow method of K-means clustering to find out highly significant up-regulated; the optimum number of clusters is 7 **b.** Cluster analysis of up-regulated genes - as K-means clustering for up-regulated is 7, the data is divided into 7 clusters. For up-regulated genes, clusters 1, 5 and 7 showed P< 0.001; 87 genes are highly up-regulated in osteophytes **c.** t-SNE plot of ‘highly differentiated’ genes (generated after K-means cluster analysis). t-SNE is a dimensional reduction technique of presenting large gene data sets; in simpler words, it represents a mathematical extract of all the selected data in the form of different variables (generally, 2, 5 or 10 variables are used). We generated a two-dimensional t-SNE plot to verify the cluster analysis. For this we considered logFC, logCPR and *p*-values of the highly significant genes in the osteophyte specimens. **d.** qRT-PCR analysis to validate transcriptome results of the osteophyte samples. mRNA levels of CPA3, CMA1, TPSAB1, MMP-1, MMP-3 and MMP-13 were normalized against the amount of ACTB. TPSAB1 (20.04-fold), CMA1 (15.57-fold), MMP-1 (16.32-fold) and MMP-3 (21.5-fold) showed a significant up-regulation. CPA3 (6.64-fold) and MMP-13 (4.01-fold) also showed up-regulation, which remained insignificant on statistical scale however. ^*^ *P* < 0.05, ^**^ *P* < 0.01

### Functional annotation of the differentially expressed gene networks

To find the functional relationship among the differentially expressed up-regulated genes, we performed a pathway enrichment analysis. Enrichment analysis is a popular method to identify biological themes in the complex lists of differentially expressed genes, using a context of prior knowledge. For this, we used Enrichr software, which contains 180184 annotated gene sets from 102 gene set libraries at present [**11**].

The enrichment analysis indicated a significant activation of matrix metalloproteinase as a top enriched canonical pathway with a combined score of 17.59 (Table 1). Additionally, osteoblast signalling (score 16.86), Receptor activator of nuclear factor kappa-B ligand (RANKL/NFκB) signalling pathway (score 16.71), angiotensin-converting enzyme (ACE) inhibitor pathway (score 16.47) and osteoclast signalling pathway (score 15.73) were the other highly enriched pathways. Inflammatory response pathway (WP453) provided an indication of immune-regulation and signalling. In brief, the functional analysis provided a footprint of bone and cartilage ECM remodelling and inflammatory responses, especially mediated by the canonical RANKL/NFκB signalling pathway.

**Table 1:** Key pathways with their *P*-values, Z-score and combined score, generated during function analysis of the differentially expressed genes in osteophyte samples.

| **Sr. No** | **Name of the Pathway (with WikiPathway No.)** | ***P*-value** | **Adjusted *P*-value** | **Z-score** | **Combined score** |
| --- | --- | --- | --- | --- | --- |
| 1 | Matrix Metalloproteinases WP129 | 0.0005743 | 0.01594 | -2.36 | 17.59 |
| 2 | Osteoblast Signaling WP322 | 0.00005487 | 0.004648 | -1.72 | 16.86 |
| 3 | RANKL/RANK (Receptor activator of NFkB (ligand)) Signaling Pathway WP2018 | 0.0002299 | 0.008505 | -1.99 | 16.71 |
| 4 | ACE Inhibitor Pathway WP554 | 0.003794 | 0.07020 | -2.95 | 16.47 |
| 5 | Osteoclast Signaling WP12 | 0.00008374 | 0.004648 | -1.68 | 15.73 |
| 6 | Photodynamic therapy-induced NF-kB survival signaling WP3617 | 0.0009077 | 0.02015 | -1.81 | 12.69 |
| 7 | Inflammatory Response Pathway WP453 | 0.01159 | 0.1040 | -2.65 | 11.80 |
| 8 | Oncostatin M Signaling Pathway WP2374 | 0.005383 | 0.07468 | -1.93 | 10.06 |
| 9 | Composition of Lipid Particles WP3601 | 0.04800 | 0.2424 | -3.27 | 9.92 |
| 10 | GABA receptor Signaling WP4159 | 0.01235 | 0.1040 | -2.18 | 9.60 |

### Up-regulated genes

Up-regulated genes in the osteophytic tissue recorded several interesting observations from OA pathological point of view. Prominently up-regulated genes were listed in Table 2. Significant up-regulation of chymase 1 (CMA1; 5-fold) carboxypeptidase A3 (CPA3; 4-fold), MS4A2/FC epsilon R1 (FCERI; 4.2-fold) and interleukin 1 receptor like 1(IL1RL1; 2.5– fold) demonstrated an active involvement of mast cells in osteophytes pathobiology. CMA1 is the only gene found in human, which encodes a chymotryptic serine proteinase. This gene is expressed in mast cells and is involved in ECM degradation. CPA3 is a common component of mast cell granules and found involved in the degradation of endogenous protein. MS4A2/FCERI is a member of membrane-spanning 4A gene family; the gene encodes a beta subunit, which is a high affinity IgE receptor and provides trigger for mast cell degranulation. IL1RL1, also known as IL-33R or ST2, is a receptor for IL-33 cytokine, which is a member of interleukin-1 family. The receptor is predominantly expressed in immune cells like mast cells and basophils.

**Table 2:**
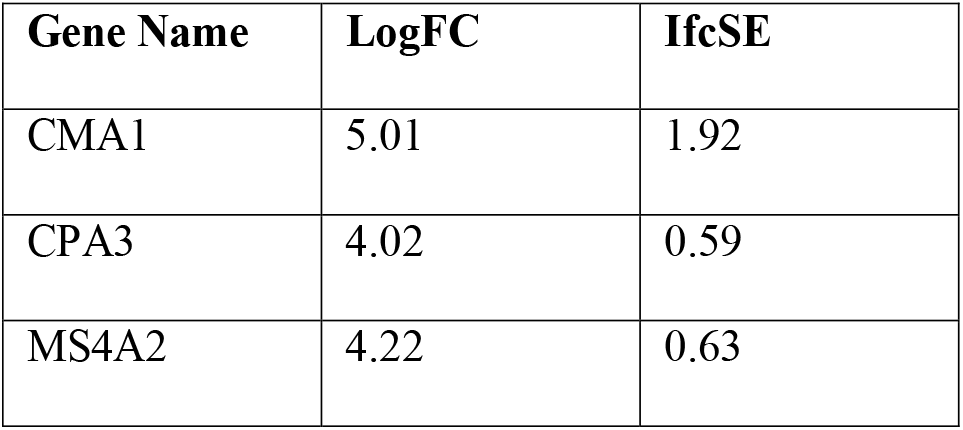
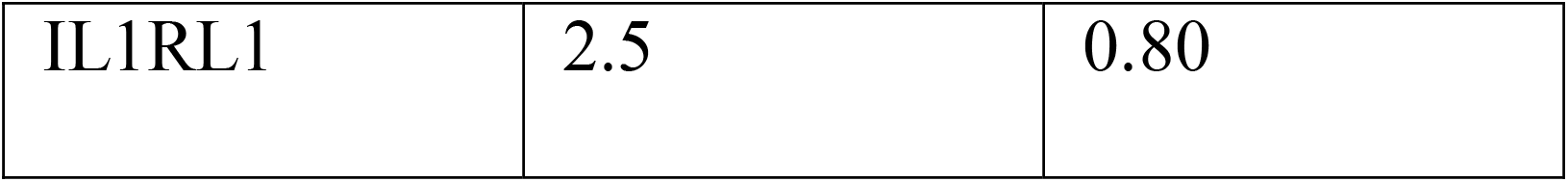
Patient to patient variation revealed by mast cell specific markers in the transcriptome analysis, wherein LogFC showed the difference of the gene expression between OA cases and controls and IfcSE was a standard deviation of the difference.

Additionally, osteophytes specimens were also characterized by a marked up-regulation of E selectin (SELE; 2.5-fold), collagen type-I alpha-1 chain (COL1A1; 2.04-fold) and COL1A2 (2.01-fold). SELE is a cell adhesion protein produced by the endothelial cells in response to cytokines and heparin and facilitates recruitment and extravasation of immune cells. COL1A1 and COL1A2 encode endochondral ossification and indicate fibrocartilageous nature of the tissue. Furthermore, matrix metalloproteinase-1 (MMP-1; 3.03-fold), MMP-3 (3.54-fold) and MMP-13 (3.2-fold), the key genes which mediate ECM remodelling, were also significantly up-regulated. To validate the transcriptomics findings, qRT-PCR was performed using CPA3, TPSAB1, CMA1, MMP-1, MMP-3 and MMP-13 genes and results are presented in **Figure 2d**.

### Immunohistochemistry (IHC) of the osteophytes

Histopathological investigation of the osteophytic tissue revealed bone matrix with distinguishable osteoblasts, osteocytes and bone-matrix interspersed with occasional blood vessels arriving from the subchondral bone as well as the cartilage with distinguished columnar chondrocytes embedded in the cartilage matrix. IHC staining with mast cells specific antibodies anti-TPSAB1 and anti-FC epsilon RI revealed an extensive presence of mast cells in all the osteophyte samples **Figure 3** (P1, P2, P3). Both antibodies-stained cells were profoundly visible in the fibrocartilage and bony tissue of the osteophytes. The cells were grouped in large number along with inner lining of osteon as well as in the space between subchondral cancellous bone trabeculae in the osteophyte samples. Moreover, the cells were found localized along with chondro-osseous junction, where the osteophytic cartilage and bone meet.

**Figure 3.**
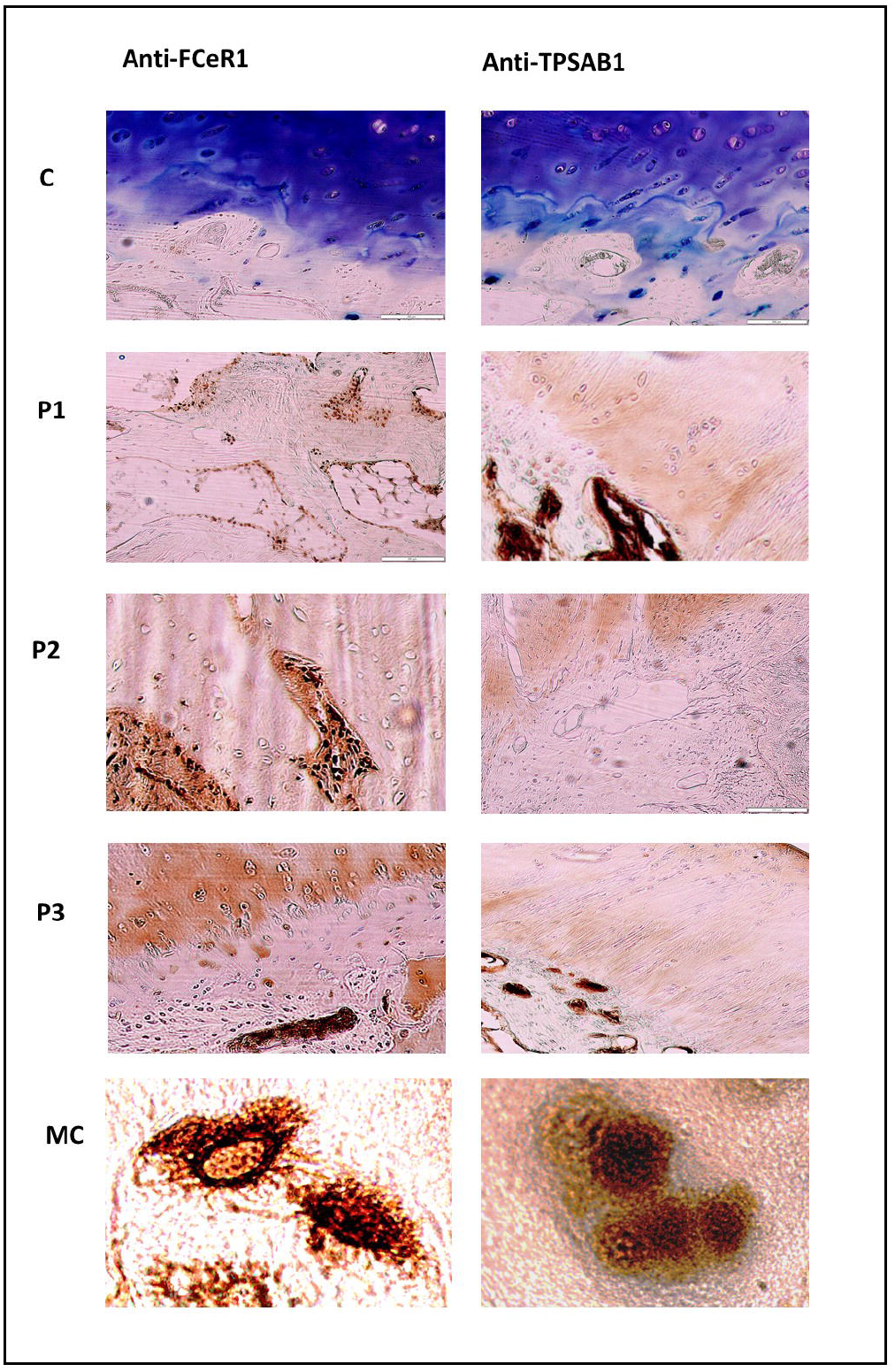
Representative slides of immunohistochemical (IHC) staining with anti-FC epsilon RI and anti-TPSAB1 to visualize mast cells in osteophyte sections. Representative areas of osteophytes of study patients are showed as P1, P2 and P3. Antibody-stained mast cells were localized in bone trabeculae, cartilage region, where the columnar chondrocytes can be seen. MC are granular mast cells at 100X magnification revealing granular feature.

Mast cell staining frequency was determined by ordinal method as described [**12**]. Anti-TPSAB1 staining incidence was comparable between control and the osteophyte samples (incidence range: 1-3%) except in one osteophyte sample (incidence range: 40%) (**Figure 3** C, P1, P2, P3-anti-TPSAB1 panel). In this osteophyte sample, mast cells were extensively gathered in the space between subchondral cancellous bone trabeculae (**Figure 3** anti-TPSAB1 panel). Unlike anti-TPSAB1, we found a significant difference for anti-FC epsilon RI staining in osteophyte samples as compared to control. While anti-FC epsilon RI staining incidence in control tissue ranged between 5-30%, it varied 10-70% for the osteophyte samples. IHC study observations corroborated with the expression signatures of the mast cells revealed in the transcriptome study.

### *In vitro* cell differentiation assays

The IHC results revealed outnumbered mast cell population in fibrocartilagenous region of the osteophyte samples. This region is directly exposed to SF. SF of progressive OA grades is known for its cytokine/chemokine composition and this innate composition was demonstrated to induce the disease specific inflammation on synovial fibroblast [**13**]. On the other hand, different lineages of immune cells circulate in their precursor form and can differentiate further into mature effector disposition after infiltration in the tissue [**14, 15, 16**]. With this existing knowledge, the authors were interested to explore the role of SF in mast cell precursors (MCPs) maturation. For this, ThP1 (human monocytes) and haematopoietic stem cells (HSCs) were treated with SF from different OA grades (10% of culture medium) and were followed for 9 days for their differentiation process. The 10^th^ day flow-cytometry analysis of the treated cells showed a clear differentiation of ThP1 cells into HLA-DR+ and CD206+ cells, while the treated HSCs were found FCERI+ phenotype (**Figure 4** – **a, b** and **c**). Outcomes of this in vitro cell differentiation assay therefore revealed that, OA SF is able to drive a clear differentiation of monocytes into macrophages and HSCs into mast cells. All the flow-cytometry experiments were performed in the triplicates and repeated for three times. Statistical analysis was carried out using GraphPad Prism 5 Software (San Diego, California, USA) using one-way ANOVA. ‘*P*’ values < 0.05 were considered as significant (**Figure 4** – **a1, b1** and **c1**).

**Figure 4.**
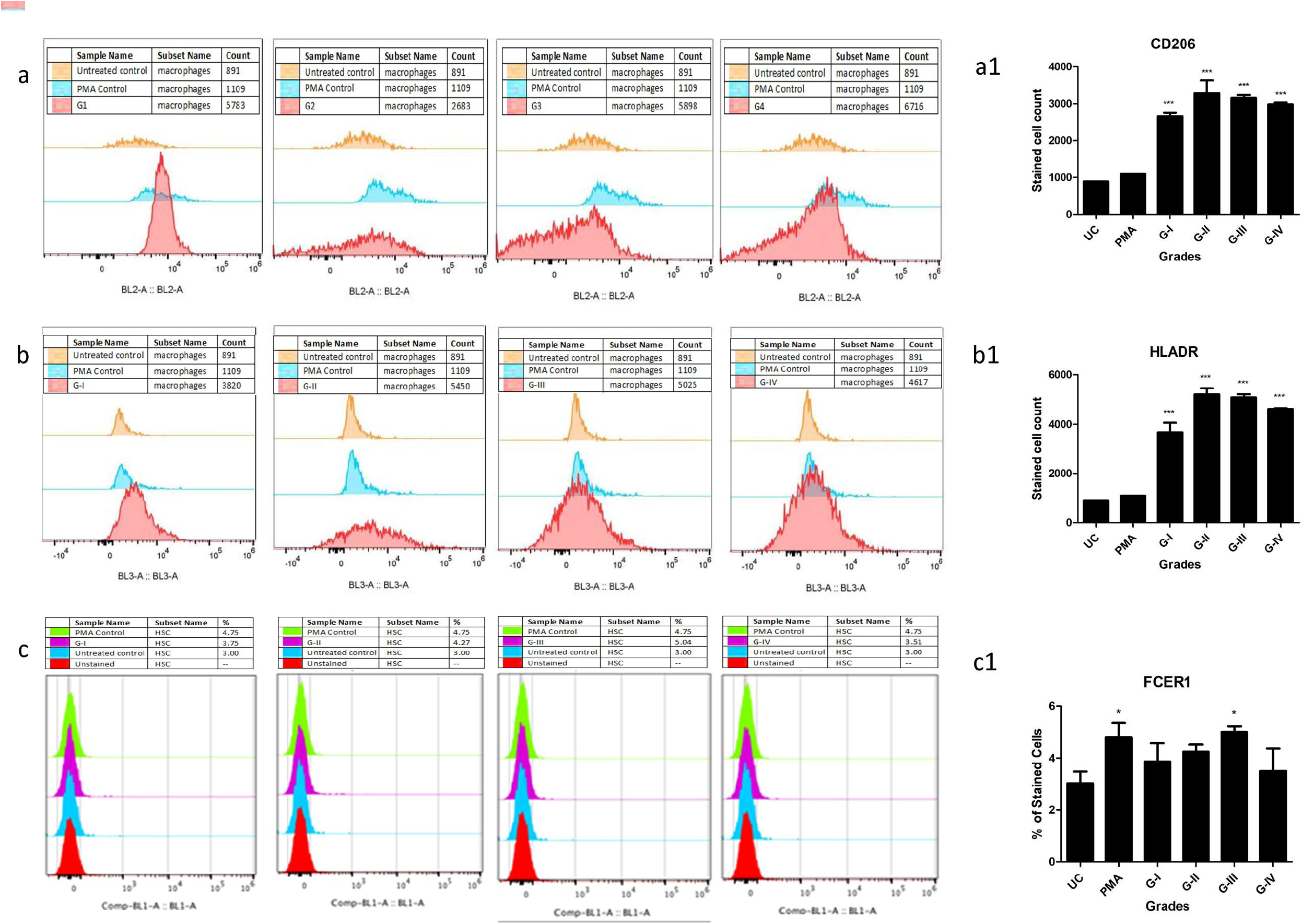
*In vitro* cell differentiation assay, where ThP1 and HSCs were treated with 10% (of culture medium) SF of grade-I to IV for 9 days; expression of cell surface markers were analysed by flow cytometry. Shown are the representative overlaid histograms from 2 independent studies, while bar graphs are the summery of the expressions. **a** – sequence of ThP1 to macrophages differentiation analysed by CD206, **b** – sequence of ThP1 to macrophages differentiation analysed by HLA-DR, **a1** and **b1** – represent summery of CD206+ and HLADR+ expression values on ThP1 to macrophages differentiation sequence; **c** – sequence of HSCs to mast cells differentiation analysed by FCERI, **c1** – represent summery of FCERI+ expression values on HSCs to mast cell differentiation sequence; ^*^ *P* < 0.05, ^**^ *P* < 0.01, ^***^ *P* < 0.001

### Proteomics analysis of OA SF

Since SF was observed to provide immunomodulatory environment for the differentiation of immune cell precursors, a proteomic investigation of OA SF was carried out. For this, OA SFs (n = 16) from different grades were subjected to LC/MS/MS analysis after depletion for high abundance proteins. The expressed proteins were identified against UniProt (www.uniprot.org) *Homo sapiens* reference proteome database. We further performed ‘paired t-test’ analysis on the grade-wise expressed proteins, where the proteins expressed in Kellgren-Lawrence radiographic (KL) grade-I SFs were used as a control group. This grade-wise comparison revealed 799 differentially up-regulated proteins as compared to KL grade-I SFs. Light chain immunoglobulins, pro-inflammatory S100 proteins, histones, actins, mitogen activated protein kinase (MAPK) family and mast cells de-granulation proteases like carboxypeptidase and cathepsins were the major subset of proteins found differentially up-regulated (Supplementary Table, **ST 2**). A grade-wise picture of these proteins and key Rectome pathway functional analysis of these protein subsets is presented in **Figure 5**. Involvement of the pathways like innate immune system, Fc-gamma receptor (FCGR) dependent phagocytosis coupled with marked accumulation of light chain immunoglobulins were potentially suggested the mast cell activation. On the other hand, differentially up-regulated proteases like carboxypeptidase, carboxypeptidase Q and cathepsins (L1, D, B and G) were the signs of mast cell function.

**Figure 5.**
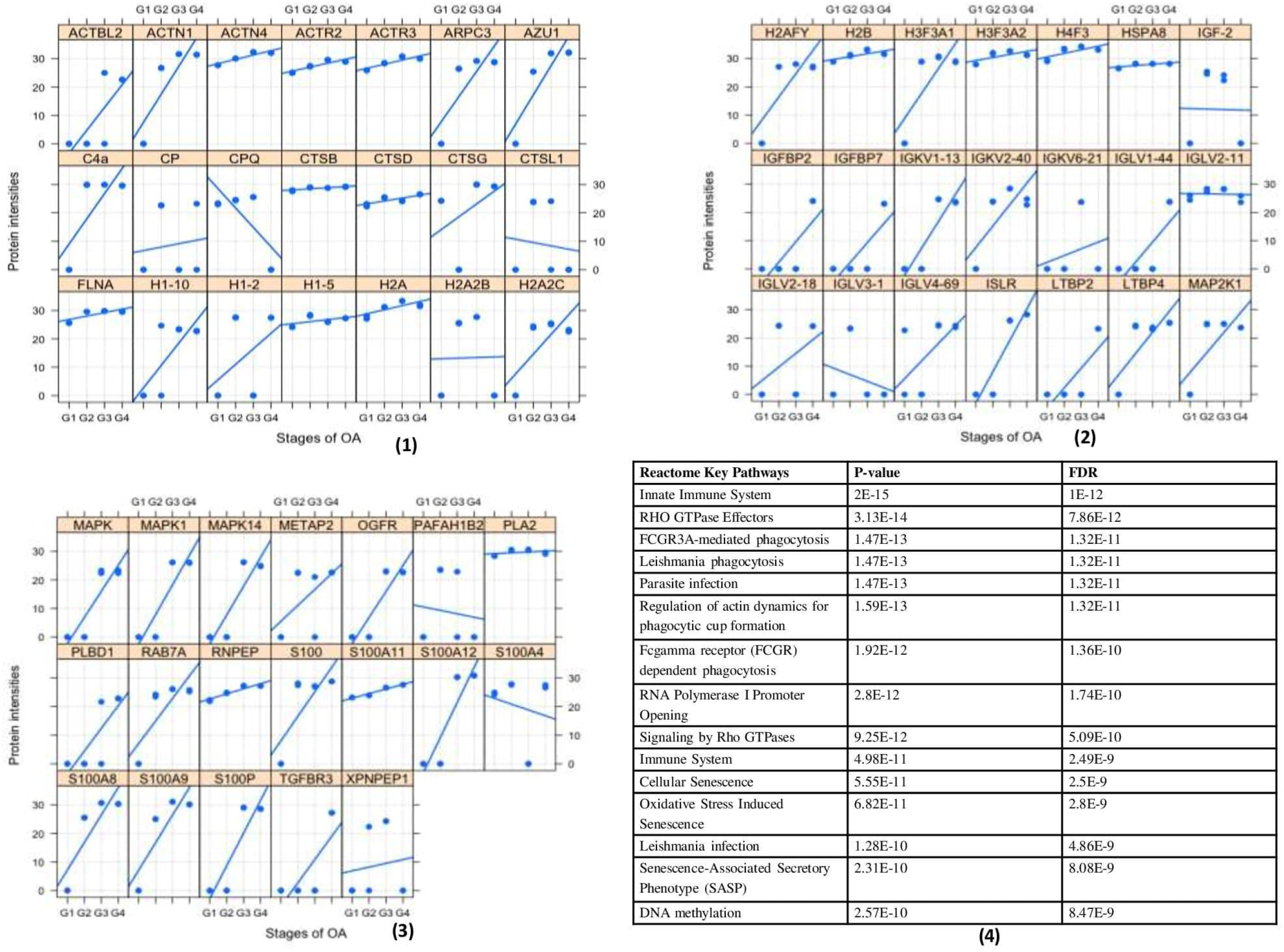
Proteomics analysis of OA SF For the proteomics analysis, four patients of each grade (4 × 4 = 16) were analyzed through MS/MS. (1), (2), and (3) represents a grade-wise picture (Lattice plots) of different protein subsets that were differentially expressed in OA SFs; differential protein expressions were determined by comparing the fold change of each protein found in KL grade-I SFs. In total, 799 proteins were found differentially expressed. These proteins contain mast cell regulatory factors like, light chain immunoglobulins, S100 A12, histones, actins, mitogen activated protein kinase (MAPK) family and mast cells de-granulation proteases like carboxypeptidase and cathepsins. (4) – functional analysis of the proteomics study in the form of key Reactome pathways with their *P*-values and FDRs

## Discussion

Although osteophytes are hallmark of OA joints, the associated molecular events are not much investigated. Transcriptome analysis showed an anticipated footprint of bone and cartilage re-modelling with significant up-regulation of MMP and osteoblast signalling pathways (**Table 1**) that involve cathepsins, COL1A1, COL1A2, MMPs and osteoglycin (OGN) transcripts. Cathepsin G (6.38-fold) and cathepsin K (2.24-fold) are among the significantly up-regulated cathepsins and are the members of cysteine proteinase pathway. Unlike the other cathepsins, cathepsin K has an ability to cleave helical and telopeptide regions of collagen 1, the major type of collagen in bone [**17**] that is best known for its active involvement in bone resorption. Expression of cathepsin K was found in osteoclasts [**18**]; moreover, along with cathepsin L and B, it was detected in the zones of active bone remodelling in osteophytes [**19**]. Cathepsin G is one of the proteases in pre-packaged granules of mast cells, which activates MMPs for ECM protein degradation [**20**]. Cathepsin G mediates release of soluble form of RANKL from the bound RANKL present on the surface of osteoblasts that leads to wider activation of osteoclasts and ultimately osteolytic lesions [**21**]. RANKL signalling pathway was found as one of the activated pathways in our analysis (**Table 1**). COL1A1 and COL1A2 are the bone matrix proteins and their expression level defines osteoblast maturation stage [**22**]. A significant up-regulation of both the genes in the osteophyte samples indicated a presence of fully mature osteoblasts. OGN (2.89-fold) is a crucial bone anabolic factor and found elevated during osteoblast differentiation in a transfection mouse model involving osteoblastic (MC3T3-E1) and myoblastic cells (C2C12) [**23**]. Our understanding on osteophyte formation process in OA is greatly influenced by Blom et al., 2004 and Gelse et al., 2012 [**3, 6**]. Growth factors, especially TGFβ and BMPs are attributed to trigger and carry growth of osteophytes, however, none of these growth factors were found up-regulated in our transcriptomics. Perhaps this is because, the osteophytes collected for this study were fully grown and likely in senescence phase as they were obtained from the patients undergoing knee replacement surgery and were unlikely to reflect any developmental changes.

MMP-1 (3.02-fold), MMP-3 (3.53-fold) and MMP-13 (3.19-fold) were found prominently up-regulated in the osteophytes and were also validated by qRT-PCR (**Figure 2d**). Role of MMPs in osteophytes is linked with multiple functions including ECM remodelling. Bone and ECM remodelling in osteophytes shows a mechanistic similarity with the process of endochondral ossification in growth plate formation. At the molecular level, ECM remodelling is carried out by the action of several MMPs, for example, proliferating chondrocytes express collagen type-II, whereas the hypertrophic chondrocytes express collagen type-X and MMP-13. On the other hand of the endochondral ossification, osteoblast express MMP-13 and collagen type-I, whereas osteoclasts express MMP-9. MMPs are directly involved in degradation of ECM components, collagenase or proteoglycan [**24**]. IHC of osteophytes samples revealed a wide spread of MMP-1 in cartilaginous tissue, fibrous tissue as well as in osteophytic osteoblasts and osteoclasts. Additionally, MMP-3 expression was reported in immature fibrous tissue of developing osteophyte and in adjacent areas of vascular invasion [**25**]. Osteoblasts, isolated from osteophytes of OA joints were showed to produce MMP-13 and further predicted to contribute in worsening of OA pathology [**26**]. Gelse et al., 2012 reported a presence of MMP-13 and MMP-9 in osteophytic cartilage [**6**]. Thus, our transcriptomics results for MMPs were in line of the previous research work.

CMA1 (5-fold), CPA3 (4.02-fold), MS4A2/FCERI (4.22-fold) and IL1RL1 (2.5-fold) were among the prominently up-regulated genes and were expressed in all the osteophyte samples of the study patients. Patient to patient variation of these genes is presented in **Table 2**. These genes are signature of mast cells and ultimately indicated their active involvement in the molecular events associated with osteophytes. MS4A2/FCERI is a primary high-affinity IgE receptor of mast cells through which IgE mediates their degranulation, whereas CMA1 and CPA3 are mast cell specific degranulation products. IL1RL1, the receptor for cytokine IL-33, belongs to toll-like receptor superfamily. IL-33 is synthesized *de novo* by the activated mast cells and is recognized as a mediator of sterile inflammation. The cytokine is found augmented in many allergic diseases as well as in rheumatoid arthritis, psoriasis and atherosclerosis. More importantly, IL-33 binding to IL1RL1/ST2 induce differentiation, survival and chemotaxis of the mast cells and further activate them to produce various cytokines [**27, 28**].

In OA, mast cells are one of the major infiltrates of inflamed synovium along with macrophages and T cells. Chou et al., 2020 identified mast cells as one of the twelve major cell types in synovium using single cell RNA sequencing [**29**]. The number of mast cells was positively correlated with the degree of synovitis and OA severity, measured by KL score [**8**]. Active participation of mast cells in OA pathology was confirmed by animal studies, wherein two distinct mast cell deficient mice model [C57BL/6J-KitW-sh/W-sh (KitW-sh/W-sh) mice and Cpa3-Cre; Mcl-1fl/fl (Hello Kitty) mice] were found protected significantly OA-related pathologies such as cartilage-loss, synovitis and osteophytes formation [**9**]. Furthermore, substantially higher CPA3 and chymase levels were detected in OA SF and their concentrations were correlated with MMP-2 and MMP-9 levels [**30**]. Presence of CPA3 was documented in osteophytic cartilage [**6**]. These evidences successfully establish involvement of mast cells in OA. Our data further marks a significant existence of mast cells in osteophyte samples and present them as an effector cells, contributing in ECM and bone remodelling process of osteophytes.

Prominent mast cell activity in the osteophyte samples can be interpreted in the form of significant up-regulation of PL2G2A and MMP-13, as the inflammatory repercussions. Number of tryptase positive mast cells were found corelated with enzymatic activity of MMP-13 in periapical lesions of inflammatory periodontitis, which were undergoing active remodelling like the osteophytes [**31**], while MMP-13 is a known indicator of inflammation in OA pathology [**32**]. In addition to the proteolytic activity of MMP-13, mast cell specific tryptase and chymase are also responsible for bone and cartilage remodelling and release cartilage and cellular degradation products, which are similar to damage associated molecular patterns (DAMPs). These cartilage and cellular breakdown products serve as endogenous agonist that interact with toll like receptor and generate a sterile inflammatory response [**33**]. Consequently, osteophytes serve as source of breakdown products that further promote worsening of OA. We previously demonstrated a temporal relationship in accumulation of glycosaminoglycan in SF with increasing severity of OA [**34**]. Significant expression of PLA2G2A (4.64-fold) in osteophyte samples can be designated as an inflammatory repercussion of mast cell activity. PLA2G2A encodes secreted phosphplipase A2, which hydrolyze Sn2 position of phospholipid molecule and usually release unsaturated fatty acids like arachidonic acid that in turn, promotes biosynthesis of inflammatory prostaglandins. PLA2G2A is often called as ‘inflammatory secretory phospholipase A2’ (sPLA2) and is involved in many inflammatory pathologies like rheumatoid arthritis and psoriasis [**35**]. Bingham III et al., 1999 reported a presence of PLA2G2A in the granules of mast cells [**36**], while Enomoto et al., 2000 demonstrated redistribution of PLA2G2A from granules of resting mast cell to the fusion sites of granules and plasma membrane in its activated form, suggesting that PLA2G2A might produce fusogenic <u>lysophospholipids</u> *in situ*, to facilitate exocytosis in bone marrow derived mast cells [**37**]. Of note, PLA2G2A is secreted by other cell types such as platelets, neutrophils, macrophages, importantly however, exogeneous PLA2G2A is able to stimulate mast cell degranulation as showed by Murakami et al., 1993 in bone marrow derived mast cells [**38**].

The transcriptome outcomes of this study indicated a link between osteophytes and mast cells; hence, we further attempted to explore the origin of the cells in osteophytes. MCPs are known to originate in bone marrow by haematopoiesis and osteophytes do have miniscule of bone marrow depending on its size and maturity. Therefore, subchondral bone is a logical route for mast cells invading into osteophytes however, there is no published evidence endorsing this channel. Also, this route does not explain how MCPs get matured into effector mast cells. Nevertheless, our IHC study observation wherein, the antibodies-stained cells were seen in noticeable number in the space between subchondral cancellous bone trabeculae in the osteophyte samples, supports this purview that MCPs from subchondral bone marrow could be a source of mast cells in osteophytes. Additionally, the authors want to claim a possibility of synovium and SF mast cells contributing to osteophytes remodelling and present the evidence to support their claim. IHC study of the osteophyte samples showed extensive accumulation of anti-FC epsilon RI and anti-TPSAB1-stained cells in cartilaginous region as compared to subchondral bone, suggesting that the mast cells invade into osteophytes through this region and reach the tide mark of cartilage-subchondral bone junction. Anti-TPSAB1-staining was also observed in zones of cartilage and subchondral bone, indicating extracellular tryptase in the tissue as a sign of degranulation of mast cells.

Further, the authors propose that invasion of the mast cells into osteophytes is mediated by SF (**Figure 6**). OA SF plays a major role in maturation of MCPs by providing necessary cocktail of cytokines, cell degradation products, and immunoglobulins. In our previous studies, we showed that this cocktail can be used to induce inflammation in HIG82, SW982, ThP1 and U937 cell lines [**13, 39, 40**]. As demonstrated by *in vitro* cell differentiation assays, differentiation of HSCs and ThP1 cells into FCERI+ and HLA-DR+/CD206+ phenotypes, respectively after SF treatment, indicate a decisive role of OA SF in differentiation of polarised cells into effector cells such as mast cells and macrophages (**Figure 4** – **a, b, c**). The results depicted in Figure 4 C should be interpreted on the basis of mast cells proportion in OA SFs as reported [41, 42, 43]. The cell differentiation can be attributed to suitable protein milieu provided by SF that contains differentially expressed mast cell regulatory proteins like, Ig light chains, free histones and S100 proteins (**ST 2**). Ig light chain proteins (kappa and lambda) have capability to bind to FCER1 and FCgR1 receptors on mast cells that ultimately trigger hypersensitive response [44]. Similarly, S100A12 at low level act as chemotactic agent for mast cells, while their high concentration trigger degranulation (**Figure 5**) [**45, 46**]. As a result of augmented inflammation and extensive cell death, MAPK proteins, free histones, and high mobility group box-1 (HMGB-1) are released in extra cellular fluids. Like the others cell degradation products, these free histones are potent inducers/activators of immune cells (**Figure 5**). Xu et al., 2009 and Xu et al., 2011 demonstrated lethal effect of intravenous injection of histone, whereas sublethal doses of histones resulted in high levels of TNF-α, IL-6, and IL-10. However, this effect was absent when TLR4 knock-out (KO) mice were used, indicating that free histones induce inflammation via TLR4 and TLR2 receptors [**47, 48**]. Therefore, it is likely that dying cells release free histones in SF of OA joints, which ultimately elevate inflammation by stimulating various immune cells including mast cells. Interestingly, our thinking is supported by Tasaka et al., 1990 wherein, exposure of rat peritoneal mast cells to a mixture of histone resulted in instant degranulation and release of histamine, indicating that mast cells readily respond to free histone [**49**]. Of note, significant elevation in MAS-related G protein-coupled receptor-X2 (MRGPRX2) (4.5-fold, in the transcriptome analysis) along with its receptor neurotensin (37.18-fold, as detected differentially expressed in SF proteomics analysis) is a strong indicator of existing IgE independent pathway of mast cell activation in osteophyte samples. MRGPRX2 binds to cationic ligands, neuropeptides and opioids and therefore represents IgE independent pathway of activation. Both the pathways trigger distinct patterns of secretion of mast cell mediators [**50**]. The outcomes from IHC and *in vitro* cell differentiation assays provide a clear indication that besides subchondral bone, an alternate route for mast cells invasion into osteophytes exists and deeper confirmation studies on this line should take place (**Figure 6**). Furthermore, significant up-regulation of SELE and chemokines like chemokine ligand-9 (CXCL9), CXCL10 and CXCL11 was likely to be associated with increased mast cell deployment into osteophytes because of mast cell specificity of these chemokines [**51**].

**Figure 6.**
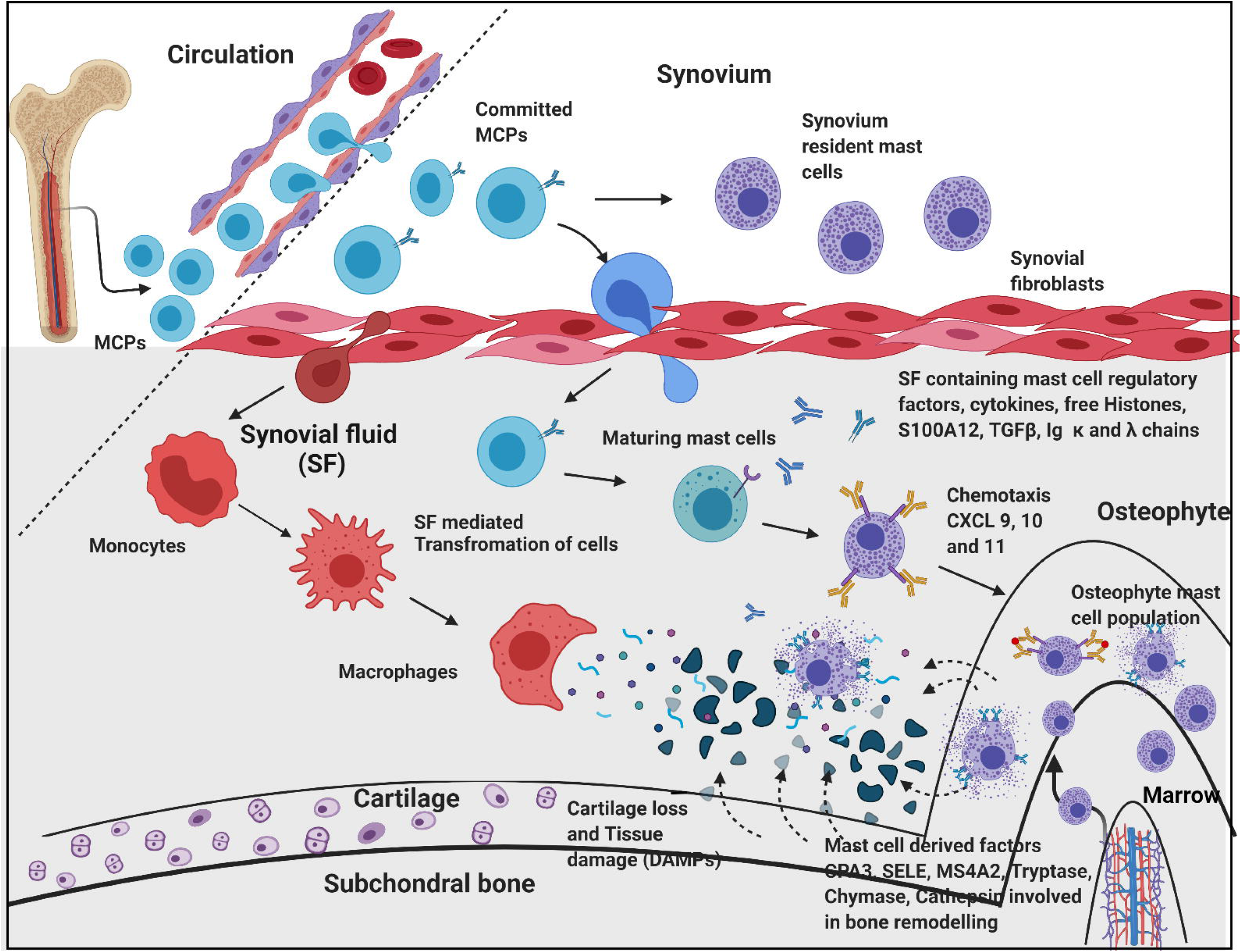
An overview of pathogenic dimension around osteophytes pertinent to mast cell action Circulatory mast cell progenitors (MCPs) are the major source of mast cells in osteophytes. OA SF, which is composed of cytokines and mast cell regulatory factors like histones, S100A12 and IgG kappa and lambda chain, play an important role in transformation of MCPs and monocytes into effector cells like mast cells and macrophages, respectively. Responding to the chemotactic signals (CXCL 9, 10 and 11) mast cells migrate and populate osteophyte. Mast cell de-granulation factors like CPA3, tryptase, chymase, cathepsins, and SELE are involved in elevated ECM remodelling of osteophyte. These degradation products and cell debris (DAMPs) accumulate in SF, which act as inducers for mast cell via TLR-2/4 pathway. Increasing action of active mast cells and macrophages contribute for synovitis, tissue damage, and cartilage-loss. This figure is created with **BioRender.com**.

Mast cells in osteophytes appear to be a new dimension of OA pathophysiology. Their invasion into cartilaginous region in a significant amount demands a deeper investigation from pathological point of view. This is because, unlike other tissues, aneural and avascular cartilage is unlikely to produce any allergic or inflammatory response and therefore raises a question about the role of allergic or inflammatory mast cells in osteophytes.

## Methods

### Statement

All methods performed in this study abide by the Declaration of Helsinki and the study protocols used in this research were approved by the Institutional Ethics Committee of University of Tartu (227/T-14 and 76785). All the recruited patients signed an informed written consent form to participate in the study.

### Osteophyte samples collection

Osteophytes were collected from six knee OA patients during knee replacement surgery. They were obtained from medial condyle of the tibia. Control tissue was non-osteophytic bony tissue obtained from lateral condyle of the tibia. Of note, the superior surface of lateral condyle of tibia is peripherally covered with fibrocartilagenous plate from where the control tissue was obtained. Thus, composition of the control tissue had a similar tissue make-up of the osteophytes, bony tissue with a sheath of fibrocartilage.

OA diagnosis was based on clinical and radiological evaluation and was performed by an experienced orthopaedic surgeon. The disease severity (or OA grading) was determined using KL score. KL score system is based on radiological signs of OA; as per this system, grade-I is doubtful narrowing of the joint space and possible, indistinguishable osteophyte. Grade-II is definite clearly identifiable osteophytes and possible narrowing of the joint space. Grade-III is with moderate multiple osteophytes with definite joint space narrowing, and grade-IV is marked with large osteophytes with marked narrowing of joint space [**34**].

### SF collection

Our research group had collected approximately 100 SF samples from progressive grades of OA patients for the research purpose by knee arthrocentesis as described [**34**]. The fluid collection protocol was approved by the Institutional Ethics committee (BVDU/MC/01, 227/T-14 and 76785). The fluid collection process was performed in minor operation theatre under strict aseptic conditions to collect SFs from early grade OA patients. The fluids from advanced grade OA were collected at the time of knee replacement surgery. A Subset of SF samples from the repository available with the research group was used for *in vitro* cell differentiation assay and proteomics study as described later in this section.

### RNA extraction

Approximately, 50 mg of the collected sample (osteophytes and control) was homogenized using liquid nitrogen and 500μl of trizol reagent (Thermo Fisher Scientific Inc., CA, USA) in mortar and pestle. The homogenate was centrifuged at 12 000 x g, 10 min, at 4°C and the supernatant was transferred into a new 2.0 ml tube and incubated for 5 min at RT. 100μl of chloroform was added into the sample, incubated it at RT for 2 min and centrifuged at 12 000 x g for 15 min at 4°C; the aqueous phase was transferred into a new 2.0 ml tube. Equal amount of freshly prepared 70% ethanol was added to the sample and transferred into Rneasy Mini spin column (Qiagen, Valencia CA, USA) as per the manufacturer’s instructions. The columns were centrifuged 8000 x g, 15 sec, at RT and the flow-through was discarded. The column was washed with 350 µl of RW1 buffer and centrifuged again 8 000 x g, 15 sec, at RT. The DNase treatment was performed applying RNase-Free DNase Set (Qiagen, Valencia CA, USA)□ for that 10µl of DNase and 70 µl of RDD buffer was mixed, transferred into the column and was incubated at RT for 30 min to digest any contaminating DNA. This step was followed by a wash with 350 µl of RW1 buffer, incubation for 5 min at RT and centrifugation 8000 x g, 15 sec at RT and then flow-through was discarded. Later, 500μl of RPE buffer was added and centrifuged 8000 x g, 15 sec, at RT□ the flow-through was discarded. RPE wash was repeated twice. Finally, the column was placed into new collection tube and centrifuged at maximum speed, at RT for 2 min to elute RNA. Then, the column was transferred into new 1.5 ml tube and 30µl of nuclease free water was transferred directly onto the column membrane, incubated at RT for 2 min and centrifuged at maximum speed, at RT for 1min. Total RNA quality was evaluated using Agilent 2100 Bioanalyzer with RNA 6000 Nano Kit (Agilent Technologies Inc., CA, USA) and the quantity was estimated using Qubit 2.0 fluorometer with RNA HS Assay Kit (Thermo Fisher Scientific Inc., CA, USA). The average RNA integrity number (RIN) of samples was between 2.8 and 7.7. The samples were stored at – 80°C until further processing.

### Whole transcriptome analysis of the osteophytes

50 ng of the total RNA was amplified by applying Ovation RNA-Seq System V2 (NuGen, Emeryville, CA, USA) after which the resulting cDNAs were used to prepare DNA fragment library with SOLiD System chemistry and barcode adaptors (Thermo Fisher Scientific Inc., CA, USA). Prior to the sequencing, all 12 libraries were labelled with different barcodes and were pooled together in equal amounts. Sequencing was performed using SOLiD 5500W platform and fragment sequencing chemistry (Thermo Fisher Scientific Inc., CA, USA).

Raw reads (75 bp) were color-space mapped to the human genome hg19 reference using Maxmapper algorithm implemented in the Lifescope software (Thermo Fisher Scientific Inc., CA, USA). Mapping to multiple locations was permitted. The quality threshold was set to 10, giving the mapping confidence more than 90. Reads with the score less than 10 were filtered out. Average mapping quality was 30. Analysis of the RNA content and gene-based annotation was done with whole transcriptome workflow. Raw sequencing data with appropriate experimental information is available in the NCBI Gene-Expression Omnibus (GEO) repository under the accession number GSE66511.

After quality control of the samples, to perform differential gene expression analysis, non-normalized raw counts were used for the EdgeR package. EdgeR is flexible tool for RNAseq data analysis to find differentially expressed genes. It performs model-based scale normalization, estimates dispersions and applies negative binomial model. Further, it implements negative binomial model fitting followed by testing procedures for determining differential expression [**52**].

In order to find differential transcriptome between osteophytes and non-osteophytic control tissue, we used group-wise comparisons, where negative binomial fitting was followed by exact test. False discovery rate (FDR) adjustment was used for multiple testing corrections. FDR threshold of 0.05 was applied for statistical significance.

### qRT-PCR analysis for the transcriptome validation

To confirm the transcriptome analysis, qRT-PCR was performed using Applied Biosystems Step One Real Time PCR System (Applied Biosystems, Foster City, CA, USA). For this, primary cells, isolated from the osteophyte samples were used. PureLink RNA Mini Kit (Ambion Inc., Invitrogen Co., Carlsbad, CA, USA) was used to isolate RNA from the cells and cDNA synthesis was performed using Superscript First-Strand Synthesis System (Invitrogen Co., Carlsbad, CA, USA) as per the manufacturer’s instructions. TaqMan gene expression assays used (Applied Biosystems, Foster City, CA, USA) to quantify the expression levels were CPA3 (gene expression assay ID - Hs00157019_m1), CMA1 (gene expression assay ID - Hs00156558_m1), TPSAB1 (gene expression ID – Hs02576518_gH), MMP1 (gene expression assay ID - Hs00899658_m1), MMP3 (gene expression assay ID - Hs00968305_m1) and MMP13 (gene expression assay ID -Hs0023392_m1). The amount of the expressed genes was normalized to the amount of ACTB (gene expression assay ID - Hs01060665_m1). Cycling conditions were 50^°^C for 2 min; 95^°^C for 10 min; and 40 cycles of 95^°^C for 15 sec and 60^°^C for 1 min. The data was analysed using Data Assist software (version 3.0).

### IHC analysis of the osteophyte samples

The osteophyte specimens were decalcified with Sakura TDE 30 Decalcifier System and were embedded in paraffin after fixation in formalin. The 5 µm sections were cut, deparaffinized and were treated with 0.9% H2O2 to inactivate endogenous peroxidase. The sections were then treated with Dako REAL Antibody Diluent (S2022; Dako Denmark A/S, Glostrup, Denmark) to block non-specific binding. After blocking, the sections were incubated with the mouse monoclonal antibody to TPSAB1(MA5-11711, Thermofisher) and rabbit polyclonal antibody to FC epsilon RI (ab229889, Abcam) to stain mast cells overnight at 4°C. Primary antibody dilution was 1:200. Visualization of the primary antibody was performed using the commercial kit “Dako REAL EnVision DetectionSystem, Peroxidase/DAB+, Rabbit/Mouse” (K5007; Dako Denmark A/S, Glostrup, Denmark). Washing steps in-between were performed using phosphate buffered saline (PBS), which contained 0.07% of Tween 20 as the detergent. Toluidine blue (Applichem, Darmstadt, Germany) was used for background staining. No immune staining was noted in negative controls, where the primary antibody was omitted. IHC images were obtained with Zeiss LSM-510 Confocal Laser -scanning microscope. Further, ordinal method was used to calculate mast cells staining frequency as described [**12**]. Ordinal method is a semiquantitative method, that reflect cellular staining frequency or intensity. Using this cellular frequency, we estimated the mast cells staining incidence (%) in the control tissues and in the osteophyte samples.

### *In vitro* cell differentiation assay

To investigate a potential of OA SF in inducing immune cell differentiation, we planned to treat ThP1 and HSCs with different grades of SF for 9 days and analyse the status of newly differentiated cells by relevant flow-cytometry markers. Here, ThP1 and HSCs were treated as immune cell precursors.

### Cell lines

#### ThP1 and HSCs

ThP1 cell line was purchased from National Centre for Cell Science (NCCS), Pune, India, while freshly isolated, certified HSCs from human bone marrow were procured from Nirav Biosolutions for this assay. Both the cell types were maintained in Roswell Park Memorial Institute (RPMI) 1640 medium (containing10% Foetal Bovine Serum (FBS) + 2 mmol/L L-glutamine + 100 U/mL *p*enicillin + 100 µg/mL streptomycin) at 95% relative humidity and 5% CO2 at 37°C.

Both the cells were seeded at a density of 1×10^5^ cells/ml and were treated with 10 % (of culture medium) SF of different OA grades. The cell differentiation was monitored for 9 days, during which a complete media change was performed every 3 days. The cells treated with phorbol 12-myristate 13-acetate (PMA) (dose – 100ng/ml) were used as a positive control, while untreated cells were used as a negative control. After 9 days of SF treatment, the adherent cells were collected for flow-cytometry analysis. To harvest the adherent cells without enzymatic digestion, the cells were incubated in 0.5 mM EDTA in Dulbecco’s Phosphate Buffered Saline for 15 minutes at 37°C and 5% CO2. After 15 minutes, the cells were collected by repeated vigorous pipetting against the bottom of the 24 well plate.

The harvested cells were fixed using (−20°C) methanol/acetone (1:1) and were blocked with human FC blocking solution (Human TrueStain FcX, Biolegends, San Diego, US) as per the manufacturer’s instruction in order to prevent any non-specific antibody binding. ThP1 cells were stained with HLA-DR (PE-Cy5.5, Miltenyi Biotec, Bergisch Gladbach, Germany) and CD206, (PE, Miltenyi Biotec, Bergisch Gladbach, Germany) macrophages specific markers as per the manufacturer’s instructions. On the other side, HSCs were stained with FCERI (FITC, Biolegends, San Diego, US) as per the manufacturer’s instructions. For each marker the median florescence intensity was measures by Attune Nxt Acoustic Flow-cytometer and graphs were plotted.

### Proteomics analysis of OA SF

Proteomics study was carried out using sixteen SFs; we included four samples of each grade (4 × 4 =16). For the depletion of high abundance albumin and immunoglobulins from SF samples, a protein depletion column was used as per the manufacturer’s instruction (BioRad proeteoMinerprotein enrichment small-capacity kit; catalogue No. 1633006). Later, protein precipitation was attained by mixing the sample with a mixture of 100% trichoroacetic acid, 0.4 % deoxycholate (TCA+DOC) in a 1:3 (v/v) ratio for 20 minutes at 4°C and later centrifuged for 15 minutes at 17000 rpm. The supernatant was discarded and in the undisturbed pellet, 3 volumes (of the original sample volume) of RT 100% acetone was added, vortexed, incubated for 10 minutes at RT and centrifuged at 17000 rpm for 15 minutes. The precipitates were air dried on ice for 10 minutes until no residual liquid was visible. Each precipitated pellet was then suspended in 100μl of 7M urea, 2M thiourea, 100mM ammonium biocarbonate (ABC) solution (7/2 urea:thiourea buffer). After reduction and alkylation of cysteine bonds with 5 mM dithiothreitol and 20 mM chloroacetamide, respectively, for 1 h at RT in the dark, the samples were digested for 4 h in 1:50 (enzyme: protein) ratio using *Lysobacter enzymogenes* (Wako Pure Chemical Industries). Solutions were diluted five times with 100 mM ABC and further digested overnight at RT with 1:50 dimethylated porcine trypsin (Sigma, Aldrich). Digested samples were then desalted using reversed phase C18 StageTips. Samples were reconstituted in 0.5% TFA for the subsequent LC/MS/MS analysis performed as described [**53**].

MS raw data were processed with the MaxQuant 1.4.0.8 software package [**54**]. Methionine oxidation, asparagine/glutamine deamidation and protein N-terminal acetylation were defined as variable modifications, while cysteine carbamidomethylation was set as a fixed modification. Peptide search was performed against in silico trypsin digested (C-terminal cleavage after lysine/arginine without proline restriction) UniProt (www.uniprot.org) *Homo sapiens* reference proteome database. First and main search MS mass tolerances were ±20 and ±4.5 ppm, respectively. MS/MS mass accuracy tolerance was ±20 ppm. Protein identifications were reported if ≥1 razor or unique peptides of ≥7 amino acids were identified. Transfer of peptide identifications (match between runs) based on accurate MS1 mass and RT was allowed. Protein quantification was reported if ≥1 peptide was quantified with ≥3 points. Label-free protein intensities were normalized using the MaxLFQ algorithm [**55**]. Peptide-spectrum match and protein false discovery rate (FDR) were kept ≤1% using a target-decoy approach. All other parameters were as default.

## Data Availability

All data produced in the present study are available upon reasonable request to the authors

## Acknowledgments

Authors acknowledge INNO-INDIGO, funding scheme for Indo-European cooperation projects in the field of research and innovation, for their financial support in the present work. We thank Dora Plus programme and Archimedes Foundation, Estonia Government for providing travel grants to PK for the research studies included in the present work.

## Contributions

A.H. S.K, and A.M. – conceived project

A.M. –sample availability, data analysis and interpretation, manuscript drafting and study supervision

P.K. – research design, performed research work, data interpretation and manuscript drafting

A.H. – research design, data analysis and interpretation, manuscript drafting and study supervision

S.K. – research design, data analysis and manuscript drafting

A.G.M. – performed research work

S.S. – performed research work

All the authors approved final draft of the manuscript

## References

1. Van der Kraan, P. M. and Van der Berg, W. B. Osteophytes: Relevance and Biology. Osteoarthr. Cartil. 15, 237–244 (2007).

2. Roelof, A. J. et al. Identification of the Skeletal Progenitor Cells Forming Osteophytes in Osteoarthritis. Ann. Rheum. Dis. 79, 1625–1634 (2020).

3. Blom, A. B. et al. Synovial Lining Macrophages Mediate Osteophytes Formation During Experimental Osteoarthritis. Osteoarthr. Cartil. 12 (8), 627–635 (2004).

4. Scharstuhl, A. et al. Inhibition of Endogenous TGF-β During Experimental Osteoarthritis Prevents Osteophyte Formation and Impairs Cartilage Repair. J. Immunol. 169 (1), 507–514 (2002).

5. Scharstuhl, A., Vitters, E. L., Van der Kraan, P. M. and Van den Berg, W. B. Reduction of Osteophyte Formation and Synovial Thickening by Adenoviral Overexpression of Transforming Growth Factor β/Bone Morphogenetic Protein Inhibitors During Experimental Osteoarthritis. Arthritis. Rheumatol. 48 (12), 3442–3451 (2003).

6. Gelse, K. et al. Molecular Differentiation Between Osteophytic and Articular Cartilage-Clues for a Transient and Permanent Chondrocyte Phenotype. Osteoarthr. Cartil. 20 (2), 162–171 (2012).

7. Brandt, K. D. Animal models of osteoarthritis. Biorheology. 39, 221–235 (2002).

8. De Lange-Brokaar, B. J. E. et al. Characterization of synovial mast cells in knee osteoarthritis: association with clinical parameters. Osteoarthr. Cartil. 24 (4), 664–671 (2016).

9. Wang, Q. et al. IgE-mediated mast cell activation promotes inflammation and cartilage destruction in osteoarthritis. eLife. 8, e39905 (2019).

10. Chen, Z. et al. The immune cell landscape in different anatomical structures of knee in osteoarthritis: a gene expression based study. BioMed. Res. Int. 2020, Article ID 9647072 (2020).

11. Kuleshov, M. V. et al. Enrichr: A Comprehensive Gene Set Enrichment Analysis Web Server 2016 Update. Nucelic. Acids. Res. 8 (44), W90–97 (2016).

12. Meyerholz, D. K. and Beck, A. P. Principles and Approaches for Reproducible Scoring of Tissue Stains in Research. Lab. Invest. 98, 844–855 (2018).

13. Koppikar, S. et al. Inflammatory response of cultured rat synoviocytes challenged with synovial fluid from osteoarthritis patients correlates with their radiographic grading: a pilot study. In. Vitro. Cell. Dev. Biol. Anim. 51 (8), 843–850 (2015).

14. Akashi, K., Traver, D., Miyamoto, T., Weissman, I. L. A clonogenic common myloid progenitor that gives rise to all myloid lineages. Nature. 404, 193–197 (2000).

15. Ginhoux, F. and Jung, S. Monocytes and macrophages: developmental pathways and tissue homeostasis. Nat. Rev. Immunol. 14, 392–404 (2014).

16. Guilliams, M., Mildner, A., Yona, S. Developmental and functional heterogeneity of monocytes. Immunity. 49 (4), 595–613 (2018).

17. Zaidi, M., Troen, B., Moonga, B.S. and Abe, E. Cathepsin K, Osteoclastic Resorption, and Osteoporosis Therapy. J. Bone Miner. Res. 16 (10), 1747–1749 (2001).

18. Zhao, Q., Jia, Y., Xiao, Y. Cathepsin K: a therapeutic target for bone diseases. Biochem. Biophys. Res. Commun. 380 (4), 721–723 (2009).

19. Lang, A., Horler, D., Baici, A. The relative importance of cystine proteases in Osteoarthritis. J. Rheumatol. 27 (8), 1970–1979 (2000).

20. Krystel-Wittemore, M., Dileepan, K. N., Wood, J. G. Mast cell: a multifunctional mast cell. Front. Immunol. 6, 620 (2016).

21. Wilson, T. J., Nannuru, K. C., Singh, R. K. Cathepsin G recruits osteoclast precursors via proteolytic activation of Protease-Activated Receptor-1. Cancer Res. 69 (7), 3188–3195 (2009).

22. Komari, T. Regulation of bone development and extracellular matrix protein genes by RUNX2. Cell. Tissue. Res. 339 (1), 189–195 (2010).

23. Tanaka, K. et al. Role of osteoglycin in the linkage between muscle and bone. J. Biol. Chem. 287 (15), 11616–11628 (2012).

24. Ortega, N., Behonick, D. J. and Werb, Z. Matrix remodeling during endochondral ossification. Trends Cell Biol. 14 (2), 86–93 (2004).

25. Bord, S., Horner, A., Hembry, R. M., Reynolds, J. J., Compston, J. E. Distribution of matrix metalloproteinases and their inhibitor, TIMP-1, in developing human osteophytic bone. J. Anat. 191, 39–48 (1997).

26. Sakao, K. et al. Osteoblasts derived from osteophytes produce interleukin-6, interleukin-8 and matrix metalloproteinase-13 in osteoarthritis. J. Bone. Miner. Metab. 27, 412 (2009).

27. Varvara, G. et al. Stimulated Mast Cells Release Inflammatory Cytokines: Potential Suppression and Therapeutical Aspects. J. Biol. Regul. Homeost. Agents. 32 (6), 1355–1360 (2018).

28. Olivera, A., Beaven, M. A. and Metcalfe, D. D. Mast Cells Signal Their Importance in Health and Disease. J. Allergy Clin. Immunol. 142 (2), 381–393 (2018).

29. Chou, C. H. et al. Synovial cell cross-talk with cartilage plays a major role in the pathogenesis of osteoarthritis. Sci. Rep. 10, 10868 (2020).

30. Zhou, X., Abdullah, N. S., Gobezie, R., Lee, D. M. and Walls, A. F. Activation of Mast Cells and their Subsets in the Synovium in Osteoarthritis (OA) and Rheumatoid Arthritis (RA). J. Allergy Clin. Immunol. Suppl.S1. 125 (2), AB178 (2010).

31. Andrade, A. L. D. L., Santos, E. M., Carmo, A. F., Freitas, R. A., Galvão, H.C. Analysis of tryptase-positive mast cells and immunoexpression of MMP-9 and MMP-13 in periapical lesions. Int. Endod. J. 50 (5), 446–454 (2017).

32. Goldring, M. B. and Otero, M. Inflammation in osteoarthritis. Curr. Opin. Rheumatol. 23 (5), 471–478 (2011).

33. Johnson, G. B., Brunn, G. J., Platt, J. L. Activation of mammalian Toll-like receptors by endogenous agonists. Crit. Rev. Immunol. 23 (1-2), 15–44 (2003).

34. Kulkarni, P. et al. Glycosaminoglycan measured from synovial fluid serves as a useful indicator for progression of osteoarthritis and complements Kellgren-Lawrence score. BBA. Clin. 6, 1–4 (2016).

35. Reddy, S. T., Winstead, M.V., Tischfield, J. A., Herschman, H. R. Analysis of the secretory phospholipase A2 that mediates prostaglandin production in mast cells. J. Biol. Chem. 272 (21), 13591–13596 (1997).

36. Bingham III, C.O. et al. Low molecular weight group IIA and group V phospholipase A2 enzymes have different intracellular locations in mouse bone marrow-derived mast cells. J. Biol. Chem. 274 (44), 31476–31484 (1999).

37. Enomoto, A. et al. Redundant and segregated functions of granule-associated heparin-binding group II subfamily of secretory phospholipases A2 in the regulation of degranulation and prostaglandin D2 synthesis in mast cells. J. Immunol. 165, 4007–4014 (2000).

38. Murakami, M., Hara, N., Kudo, I., Inoue, K. Triggering of degranulation in mast cells by exogenous type II phospholipase A2. J. Immunol. 151, 5675–5684 (1993).

39. Ingale, D. et al. Synovium-synovial fluid axis in osteoarthritis pathology: a key regulator of the cartilage degradation process. Genes. 12 (7), 989 (2021).

40. Kulkarni, P., Martson, A., Vidya, R., Chitnavis, S., Harsulkar, A. Pathophysiological landscape of osteoarthritis. Adv. Clin. Chem.100, 37–90 (2021).

41. Kriegova, E. et al. Gender-related differences observed among immune cells in synovial fluid in knee osteoarthritis. Osteoarthr. Cartil. 26 (9), 1247–1256 (2018).

42. Dean, J., Hoyland, J. A., Denton, J., Donn, R. P., Freemont, A. J. Mast cells in the synovium and synovial fluid in osteoarthritis. Rheumatology. 32, 671–675 (1993).

43. Malon, D. G., Irani, A. M., Schwartz, L. B., Barrett, K. E., Metcalfe, D. D. Mast cell numbers and histamine levels in synovial fluids from patients with diverse arthritides. Arthritis Rheum. 29 (8), 956–963 (1986).

44. Redegeld, F. A. et al. Immunoglobulin-free light chains elicit immediate hypersensitivity-like responses Nat. Med. 8, 694–701 (2002).

45. Yan, W. X. et al. Mast cell and monocyte recruitment by S100A12 and its hinge domain. J. Biol. Chem. 283, 13035–43 (2008).

46. Yang, Z. et al. S100A12 provokes mast cell activation: a potential amplification pathway in asthma and innate immunity. J. Allergy. Clin. Immunol. 119, 106–14 (2007).

47. Xu, J. et al. Extracellular histones are major mediators of death in sepsis. Nat Med. 15, 1318–1321 (2009).

48. Xu, J., Zhang, X., Monestier, M., Esmon, N. L. and Esmon, C.T. Extracellular Histones Are Mediators of Death through TLR2 and TLR4 in Mouse Fatal Liver Injury. J. Immunol. 187 (5), 2626–2631 (2011).

49. Tasaka, K., Mio, M., Akagi, M. and Saito T. Histamine Released Induced by Histone and Related Morphological Changes in Mast Cells. Agents Actions. 30, 114–117 (1990).

50. Meixiong, J. et al. Activation of Mast-Cell-Expressed Mas-Related G-Protein-Coupled Receptors Drives Non-histaminergic Itch. Immunity. 50 (5), 1163-1171.e5 (2019).

51. Ruschpler, P. et al. High CXCR3 expression in synovial mast cells associated with CXCL9 and CXCL10 expression in inflammatory synovial tissues of patients with rheumatoid arthritis. Arthritis. Res. Ther. 5, R241–R252 (2003).

52. Robinson, M. D., MaCarthy, D. J., Smyth, G. K. edgeR: a Bioconductor package for differential expression analysis of digital gene expression data. Bioinformatics. 26, 139–140 (2010).

53. Mets, T. et al. Fragmentation of Escherichia coli mRNA by MazF and MqsR. Biochimie. 156, 79–91 (2019).

54. Cox, J., Mann, M. MaxQuant enables high peptide identification rates, individualized p.p.b. – range mass accuracies and proteome-wide protein quantification. Nat. Biotechnol. 26, 1367–1372 (2008).

55. Cox, J. et al. Accurate proteome-wide label-free quantification by delayed normalization and maximal peptide ratio extraction, termed MaxLFQ. Mol. Cell. Proteomics. 13, 2513–2526 (2014).

56. Kormelink, T. G. et al. Mast cell degranulation is accompanied by the release of a selective subset of extracellular vesicles that contains mast cell-specific proteases. J. Immunol. 197, 3382–3392 (2016).

