## Supplementary Table 1 (ST1) for "Mast cells differentiated in synovial fluid and resident in osteophytes exalt the inflammatory pathology of osteoarthritis"

**Supplementary table 1 (ST1): A list of significantly up-regulated and down-regulated genes in the osteophyte samples with their LogFC and *P*-values**

| **Up-regulated Genes** | | |
| --- | --- | --- |
| **Gene Name** | **LogFC** | ***P*-value** |
| CPA3 | 4.027533 | 5.09E-18 |
| SELE | 2.536014 | 2.14E-16 |
| MS4A2 | 4.220766 | 3.78E-15 |
| PLA2G2A | 4.644088 | 5.87E-12 |
| CSN1S1 | 5.942081 | 1.44E-10 |
| HAPLN1 | 4.354046 | 1.48E-10 |
| GABRA4 | 3.303431 | 4.35E-10 |
| PRG4 | 2.714763 | 5.14E-10 |
| THBS4 | 2.993005 | 6.09E-10 |
| IBSP | 2.006641 | 7.31E-10 |
| SLC36A2 | 2.247509 | 7.09E-09 |
| HPGD | 3.213818 | 9.49E-09 |
| OGN | 2.890213 | 4.86E-08 |
| ASPN | 2.432588 | 6.50E-08 |
| CTSG | 6.380478 | 7.17E-08 |
| F5 | 2.485848 | 1.88E-07 |
| FAM38B | 2.125555 | 4.49E-07 |
| IL1RL1 | 2.520738 | 8.72E-07 |
| ZIC1 | 4.85166 | 1.12E-06 |
| PRSS35 | 2.469275 | 1.73E-06 |
| STMN2 | 2.835148 | 1.70E-06 |
| MMP-13 | 3.19846 | 2.01E-06 |
| CRTAC1 | 2.740265 | 2.06E-06 |
| SHOX2 | 2.027773 | 7.05E-06 |
| ACP5 | 2.83225 | 7.15E-06 |
| COMP | 3.324406 | 9.58E-06 |
| COL1A2 | 2.01073 | 1.29E-05 |
| LRFN5 | 3.088649 | 1.65E-05 |
| TMEM196 | 2.776174 | 1.94E-05 |
| HBA2 | 2.960499 | 2.08E-05 |
| CTSK | 2.240189 | 2.15E-05 |
| CKB | 2.25553 | 2.41E-05 |
| GNG4 | 2.377001 | 3.22E-05 |
| ADCYAP1 | 2.582899 | 3.33E-05 |
| CNR1 | 2.254559 | 5.24E-05 |
| ST18 | 2.889322 | 5.83E-05 |
| MMP3 | 3.539804 | 6.33E-05 |
| FZD10 | 2.231317 | 0.000133 |
| GJB2 | 3.455437 | 0.000135 |
| CYP27C1 | 4.37209 | 0.000173 |
| C1orf186 | 3.272109 | 0.000199 |
| HDC | 2.110747 | 0.000243 |
| CMA1 | 5.007723 | 0.000274 |
| COL1A1 | 2.038679 | 0.000368 |
| SYT6 | 2.129815 | 0.000422 |
| AMPH | 2.072226 | 0.000523 |
| AMTN | 3.814246 | 0.000729 |
| TNFSF11 | 2.405489 | 0.001057 |
| GALNT14 | 4.471982 | 0.001105 |
| TRIM11 | 2.051431 | 0.001203 |
| NELL1 | 4.426061 | 0.001209 |
| TIMD4 | 2.79137 | 0.001212 |
| HHIP | 3.320885 | 0.001394 |
| MRGPRX2 | 4.517302 | 0.001406 |
| GFPT2 | 2.213461 | 0.001616 |
| NPHS1 | 3.499433 | 0.001877 |
| RASL12 | 2.011284 | 0.002044 |
| HEMGN | 2.173148 | 0.002365 |
| CXCL9 | 2.226117 | 0.002528 |
| IDO1 | 3.382825 | 0.002929 |
| MYO3A | 2.28746 | 0.00321 |
| ADAM23 | 3.193211 | 0.003252 |
| LRRC8E | 2.234257 | 0.003983 |
| SERPINA5 | 2.802678 | 0.004112 |
| GYPA | 2.738816 | 0.004161 |
| SLC4A1 | 4.885617 | 0.004197 |
| LIF | 3.908902 | 0.004473 |
| OPCML | 2.428177 | 0.004786 |
| PART1 | 2.41668 | 0.004784 |
| CXCL10 | 2.225288 | 0.006315 |
| FAM40B | 2.022329 | 0.00656 |
| C21orf37 | 4.084623 | 0.006829 |
| WFDC1 | 2.136605 | 0.006992 |
| KIF4A | 2.248998 | 0.007723 |
| FAM133A | 2.053699 | 0.008372 |
| CLDN10 | 2.516074 | 0.009417 |
| SLC9A2 | 2.165385 | 0.010866 |
| TUBA8 | 2.401238 | 0.010872 |
| CXCL11 | 2.190709 | 0.013103 |
| SULT1B1 | 3.152541 | 0.013765 |
| MMP1 | 3.027436 | 0.014302 |
| CILP | 2.148754 | 0.014409 |
| CRLF1 | 3.817562 | 0.015141 |
| CA1 | 2.451945 | 0.017428 |
| MARCO | 3.779762 | 0.018039 |
| LINGO1 | 2.318577 | 0.018117 |
| C10orf105 | 2.176865 | 0.0222 |

| **Down-regulated Genes** | | |
| --- | --- | --- |
| **Gene Name** | **LogFC** | **P-value** |
| APOB | -2.03918 | 9.05E-11 |
| CADM2 | -2.95074 | 1.41E-08 |
| TMEFF2 | -4.15435 | 0.000413 |
| GNAZ | -3.41417 | 0.000625 |
| GABRA2 | -2.77263 | 0.000654 |
| NRG3 | -4.14112 | 0.001502 |
| CHIT1 | -3.25043 | 0.003048 |
| C6orf10 | -3.72459 | 0.003143 |
| NETO1 | -2.91119 | 0.00346 |
| FAM86DP | -3.6999 | 0.00405 |
| LOC100129345 | -3.6983 | 0.004298 |
| GLIS1 | -3.69735 | 0.004474 |
| IGSF1 | -2.84473 | 0.004914 |
| TRAF2 | -3.68328 | 0.006547 |
| SERPINA12 | -2.98392 | 0.007455 |
| TINAGL1 | -3.55897 | 0.007825 |
| LOC148824 | -3.56945 | 0.009136 |
| C1orf74 | -3.55768 | 0.009161 |
| FLJ30403 | -3.40428 | 0.015644 |
| LOC653113 | -3.43474 | 0.018687 |
| PIAS4 | -3.40745 | 0.020407 |
| PRINS | -3.40161 | 0.021045 |

**ST2:** A Grade-wise demonstration of key mast-related proteins found in the proteomics analysis of SF

| **Protein Name** | **Protein Abbreviation** | **LogFC**  **G2G1** | **LogFC**  **G3G1** | **LogFC**  **G4G1** | **Ref** |
| --- | --- | --- | --- | --- | --- |
| Immunoglobulin lambda variable 2-18 | IGLV2-18 | 37.5896545 | NA | 37.42546 |  |
| Immunoglobulin lambda variable 3-1 | IGLV3-1 | 36.59979569 | NA | NA |  |
| Immunoglobulin kappa variable 2-40 | IGKV2-40 | 37.05100621 | 41.67871 | 37.29651 |  |
| Immunoglobulin lambda variable 2-11 | IGLV2-11 | 2.431556072 | 2.782218 | NA |  |
| Immunoglobulin superfamily containing leucine-rich repeat protein | ISLR | NA | 39.41972 | NA |  |
| Immunoglobulin kappa variable 1-13 | IGKV1-13 | NA | 37.90953 | NA |  |
| Immunoglobulin kappa variable 6-21 | IGKV6-21 | NA | 36.9165 | NA |  |
| Immunoglobulin lambda variable 4-69 | IGLV4-69 | NA | 2.773235 | NA |  |
| Immunoglobulin superfamily containing leucine-rich repeat protein | ISLR | NA | NA | 41.52395 |  |
| Immunoglobulin lambda variable 1-44 | IGLV1-44 | NA | NA | 36.97934 |  |
| Immunoglobulin kappa variable 1-13 | IGKV1-13 | NA | NA | 36.86832 |  |
| Immunoglobulin lambda variable 4-69 | IGLV4-69 | NA | NA | 2.380682 |  |
| Protein S100 (Fragment) | S100A6 | 41.16882 | 40.40297 | 42.1267 |  |
| Protein S100-A8 | S100A8 | 38.80698 | 43.95978 | 43.62165 |  |
| Protein S100-A9 | S100A9 | 38.33136 | 44.35842 | 43.4431 |  |
| Protein S100-A4 | S100A4 | 3.127559 | -37.9506 | 2.521137 |  |
| Protein S100-A11 | S100A11 | 0.747533 | 3.450775 | 4.473638 |  |
| Protein S100-A12 | S100A12 | NA | 43.6355 | 44.17231 |  |
| Protein S100-P | S100P | NA | 42.32842 | 41.86717 |  |
| Histone H3.1 | H3C1 | 42.15141 | 43.86723 | 42.13659 |  |
| Histone H1.2 | H1-2 | 40.8021 | NA | 40.73825 |  |
| Isoform 1 of Core histone macro-H2A.1 | MACROH2A1 | 40.35853 | 41.30319 | 40.28084 |  |
| Histone H2A type 2-B | H2AC21 | 38.82677 | 40.98707 | NA |  |
| Histone H2A type 2-C | H2AC20 | 37.47739 | 38.57285 | 36.1769 |  |
| Histone H1.10 | H1-10 | 36.92155 | 36.63726 | 36.05325 |  |
| Histone H1.5 | H1-5 | 4.143112 | 1.894718 | 3.143245 |  |
| Histone H4 | H4C1 | 4.135902 | 5.062983 | 3.918993 |  |
| Histone H3.2 | H3C15 | 3.952981 | 4.763897 | 3.338169 |  |
| Histone H2A | H2AZ2 | 3.513314 | 38.20615 | 4.071692 |  |
| Histone H2B | H2BC15 | 2.290555 | 4.26651 | 2.707146 |  |
| Methionine aminopeptidase 2 | METAP2 | 35.78272 | 33.33631 | 35.87134 |  |
| Xaa-Pro aminopeptidase 1 | XPNPEP1 | 34.63523 | 37.59244 | NA |  |
| Aminopeptidase B | RNPEP | 2.703409 | 5.132456 | 5.127756 |  |
| Alpha-actinin-1 | ACTN1 | 40.01584 | 44.90512 | 44.6893 |  |
| Actin-related protein 2/3 complex subunit 3 | ARPC3 | 39.73583 | 42.46596 | 42.05718 |  |
| Actin-related protein 3 | ACTR3 | 2.494164 | 4.897881 | 4.0529 |  |
| Alpha-actinin-4 | ACTN4 | 2.359276 | 4.658768 | 4.330454 |  |
| Actin-related protein 2 | ACTR2 | 2.279216 | 4.53598 | 3.858559 |  |
| Actin, cytoplasmic 1 | ACTB | 2.240241 | -37.5188 | 2.776209 | [56] |
| Beta-actin-like protein 2 | ACTBL2 | NA | 37.29973 | 35.91144 | [56] |
| Platelet-activating factor acetylhydrolase IB subunit beta (Fragment) | PAFAH1B2 | 36.83481 | 35.17532 | NA |  |
| Ras-related protein Rab-7a | RAB7A | 37.22036 | 39.44037 | 38.90048 | [56] |
| Heat shock cognate 71 kDa protein | HSPA8 | 1.609628 | 1.574592 | 1.596362 | [56] |
| Filamin-A | FLNA | 3.974561 | 4.215035 | 3.994411 |  |
| Carboxypeptidase | CTSA | 35.9543 | NA | 35.5287 |  |
| Carboxypeptidase Q | CPQ | 1.294259 | 2.359289 | -36.5255 |  |
| Cathepsin L1 | CTSL | 37.15384 | 36.42123 | NA |  |
| Cathepsin D | CTSD | 2.680759 | 1.436841 | 3.760144 |  |
| Cathepsin G | CTSG | -37.5698 | 5.721166 | 5.100312 |  |
| Dual specificity mitogen-activated protein kinase kinase 1 | MAP2K1 | 38.16372 | 38.24167 | 36.92404 |  |
| Mitogen-activated protein kinase 1 | MAPK14 | NA | 39.43511 | 39.30365 |  |
| Mitogen-activated protein kinase 14 | MAPK14 | NA | 39.48305 | 38.15754 |  |
| Mitogen-activated protein kinase | MAPK3 | NA | 36.16989 | 36.2431 |  |
| Phospholipase A2 | PLA2G2A | 2.003637 | 2.129905 | 0.998775 |  |
